# Household Air Pollution Exposures Over Pregnancy and Maternal Blood Pressure Trajectories through 8 Years Postpartum: Evidence from the Ghana Randomized Air Pollution and Health Study (GRAPHS)

**DOI:** 10.1101/2025.01.17.25320752

**Authors:** Seyram Kaali, Michelle Li, Mohamed Nuhu Mujtaba, Elena Colicino, Sule Awuni, Blair Wylie, Musah Osei, Kholiswa Tsotetsi, Tawfiq Yussif, Steve Chillrud, Darby Jack, Kwaku Poku Asante, Alison Lee

**Author notes:** **Corresponding Author**: Alison G. Lee, MD MS 1468 Madison Avenue Icahn School of Medicine at Mount Sinai, New York, NY, USA., (P), (F). **Co-corresponding author**: Seyram Kaali, MD, MPH, Kintampo Health Research Centre, Research and Development Division, Ghana Health Service.

## Abstract

**Background:** Household air pollution is a major contributor to cardiovascular disease burden in women in Sub-Saharan Africa. However, little is known about exposures during pregnancy or the effect of clean cooking interventions on postpartum blood pressure trajectories.

**Methods:** The Ghana Randomized Air Pollution and Health Study (GRAPHS) randomized 1414 non-smoking women in the first and second trimesters to liquefied petroleum gas (LPG) or improved biomass stoves – vs control (traditional three-stone open fire). Personal exposure to carbon monoxide was measured at four prenatal timepoints and three times over the first postpartum year. Participants were prospectively followed with annual resting BP measurements at 2, 4, 5, 6, 7, and 8 years postpartum. We employed linear mixed effects models to determine effect of GRAPHS interventions on postpartum BP, and to examine associations between prenatal and postnatal CO and postpartum BP.

**Results:** LPG intervention was associated with 3.54mmHg (95% CI-5.55,-1.53) lower change in systolic BP from enrolment through 8 years postpartum, and 2.27mmHg (95% CI-3.61,-0.93) lower change in diastolic BP from enrolment through 8 years postpartum, as compared to control. In exposure-response analysis, average prenatal CO was positively associated with change in systolic BP from enrolment (β=0.71mmHg, 95% CI 0.08, 1.30, per doubling of CO)

**Conclusions:** LPG cookstove intervention initiated in early pregnancy and maintained through the first postpartum year was associated with lower systolic and diastolic BP trajectories through 8 years postpartum. These findings support the need to integrate clean cooking solutions into existing antenatal care packages.

## Introduction

Cardiovascular disease (CVD) is the leading cause of death in women globally, accounting for 9 million deaths and over 170 million disability adjusted life years (DALYs) in 2021(1).

Globally, hypertension, defined as blood pressure (BP) of 140/90 mmHg or higher, is one of the leading risk factors for CVD. Approximately 78% of all hypertension cases reside in low and middle-income countries (LMICs)(2). Within LMICs, sub-Saharan Africa is one of the highest burden regions, with 48% hypertension prevalence in women(3). In Ghana, hypertension ranks among the top four risk factors for death and disability(4). Compared to age-matched men, women have higher BP trajectories beginning as early as 30 years(5). Female-specific risk factors including those related to pregnancy and reproduction(6–8) may account for the sex differences in BP trajectories(5, 9). We therefore hypothesize that pregnancy is a sensitive window where lifetime CVD risk may be determined. A better understanding of modifiable risk factors during pregnancy including air pollution, and how these influence BP in the postpartum period, particularly in LMICs, is urgently needed.

Extensive research links ambient air pollution exposure to CVD risk, including BP (10–12). In LMICs, household air pollution (HAP) resulting from inefficient combustion of biomass fuels such as firewood, charcoal, animal dung, and agricultural residues in open fires or inefficient stoves is a major source of air pollution exposure. An estimated 49% of the world’s population (approximately 3.8 billion people) mostly in LMICs are exposed daily to high concentrations of HAP (9). HAP exposure is responsible for substantial morbidity and mortality, with an attributable 2.3 million deaths and 91.5 million disability-adjusted life years (DALYs) in 2019. CVD is the largest cause of HAP attributed mortality(13). In Ghana, air pollution is the second leading risk factor driving mortality and disability(4). As primary household cooks, women of reproductive age are highly exposed to HAP, but little is known about how these exposures affect BP.

Randomized cook stove intervention trials to reduce HAP exposure have enrolled pregnant women and examined associations with BP however findings are mixed. A study in Nigeria randomized pregnant women to an ethanol cookstove (n=162) versus to kerosene or firewood fuel as controls (n=162) and found that while systolic BP (SBP) was not affected, the mean diastolic BP (DBP) was on average 2.8mmHg lower in the intervention arm compared to the control group over six antenatal visits (14). Conversely, the Household Air Pollution Intervention Network (HAPIN) study randomised n=3002 pregnant women to an LPG cookstove intervention (n=1500) versus solid fuel controls (n=1502) and found that SBP, DBP and mean arterial pressure (MAP) over pregnancy were higher in the intervention arm as compared to control(15). A cross-sectional study in India of 1369 pregnant women found that compared to gas users, women using wood had on average both lower SBP (−2.0 mm Hg [95% CI, −3.77, −0.31]) and DBP (−1.96 mm Hg [95% CI, −3.60, −0.30]) and the risk of hypertension was not significantly different between wood and gas users at 24 hours post-delivery(16). A consistent limitation amongst these studies is the lack of long-term follow-up to understand how a stove intervention to reduce HAP exposures over pregnancy influence postpartum BP trajectories. In the ambient air pollution literature, prenatal air pollution has been associated with higher gestational blood pressure and increased risk for hypertensive disorders of pregnancy (17–20), however, to our knowledge no study has examined associations with postpartum BP.

To address these research gaps, we leveraged the Ghana Randomized Air Pollution and Health Study (GRAPHS) cohort. GRAPHS was a cluster-randomized cook stove intervention trial that randomised n=1,414 pregnant women to one of two cookstove interventions, a liquefied petroleum gas (LPG) stove, or improved biomass stove, or control (continued open wood fire cooking) with repeated maternal personal HAP exposure measurements over pregnancy and the first year postpartum, as indexed by carbon monoxide (CO) (21, 22). As previously published, the GRAPHS LPG intervention resulted in a 47% reduction in CO exposure over pregnancy as compared to control; no difference in exposure between the improved biomass and controls arms was observed(23). In a subset of n= 699 women, annual resting BP measurements were performed through eight years postpartum. We first examined whether either GRAPHS stove intervention was associated with BP over 8 years postpartum and whether the effects differed by fetal sex. We then examined exposure-response associations between prenatal and postnatal HAP exposure and BP and again effect modification by fetal sex.

## Methods

### Study Participants

GRAPHS was a cluster-randomized controlled trial to investigate the impact of prenatally introduced stoves to reduce HAP exposure on birth weight and pneumonia risk over the first year of life. GRAPHS has been described elsewhere (15,16). Briefly, between August 2013 and June 2015 we recruited 1,414 non-smoking pregnant women in communities in the Kintampo North and South Districts of central Ghana.

Eligible women had to be the head cook of their household with an ultrasound confirmed singleton pregnancy of less than 24 weeks gestation(24). Women in the intervention arms received either a liquefied petroleum gas (LPG) stove or an improved biomass stove (BioLite Home Stove, BioLite Inc., Brooklyn, NY). In 2017, additional funding was obtained to prospectively follow a subset of 699 GRAPHS mothers with continued exposure and health phenotyping assessments, including annual resting BP measurements through 8 years postpartum. This analysis includes n=690 women without a prior history of hypertension and who completed at least one valid BP measure. All procedures were approved by the ethics committees at all participating institutions, including the Kintampo Health Research Centre, Ghana Health Service, Columbia University, and Icahn School of Medicine at Mount Sinai, with all participants providing written informed consent.

### GRAPHS Stove Interventions

Communities were randomized to one of two cookstove interventions (LPG or Improved biomass) or open-fire control. Following participant enrollment, a baseline personal exposure measurement was performed and then the stove intervention deployed and supported for the remainder of pregnancy and through one year postpartum. Participants in the LPG arm received a two-burner LPG stove and two 14.5-kg cylinders, with monthly refill deliveries and as needed. Women enrolled in the improved biomass stove arm received two BioLite stoves which allowed the continued use of solid fuels but enhanced heat transfer and combustion efficiency by using a thermoelectric powered fan to circulate air through the combustion chamber(25). Women in the control arm continued to cook on traditional open-fire stoves. Trained fieldworkers conducted weekly household visits to all participants, encouraging intervention use and repairing the stoves as needed. All study households were provided with a treated mosquito bed net and health insurance.

Support for GRAPHS intervention ended when the index child turned one year of age. Upon GRAPHS completion at child age one year, the control and improved biomass arm households received a two-burner LPG stove and two 14.5-kg cylinders however no fuel was supplied.

### Exposure Assessment

As previously described in detail elsewhere(21, 23), maternal HAP exposure was indexed by 72-hour personal CO exposure assessments and was performed at four (4) time points over pregnancy; once prior to stove deployment, three times between stove deployment and ultrasound estimated date of delivery, and three times over the first year postpartum using the Lascar EL-CO-USB Data Logger (Lascar Electronics, Erie, Pennsylvania, USA). The Lascar monitors measured CO exposure in parts per million (ppm) every 10 s and were affixed to participant’s clothing close to their breathing zone. Participants were asked to wear the device except while sleeping and bathing, at which time they were asked to keep the monitors nearby (<1 m away) and off the floor. Time-weighted averages of CO across gestation and the first year postpartum, considered separately, were generated by linear interpolation of CO values(26). Quality control has been described elsewhere and included time check runs and visual inspection of each deployment as per protocol as well as interval exposure of the Lascar monitors to a certified span gas (50 ppm CO in zero air) at the KHRC laboratory to quantify response and adjust field values(23). As with other GRAPHS analyses, exposure data was restricted to the first 48 hours of each 72-hour measurement and only measurements that passed all quality control checks were used(27–31).

### Blood Pressure Measurements

On GRAPHS enrolment, trained fieldworkers measured maternal resting BP once using the Watch Home automated BP monitor (Microlife AG Swiss Corporation, Widnau, Switzerland). At 2 (subset of n=336), 4, 5, 6, 7, and 8 years following delivery of the study index child (hereafter referred to as postpartum), maternal resting BP was measured by trained fieldworkers using the oscillometric and digital Omron BP742N BP monitor (OMRON Healthcare, CA, USA) per protocol. Following a 10-minute period of seated rest, trained fieldworkers measured two to three systolic and diastolic BP, each 5 minutes apart using an appropriately fitted BP cuff. If three measurements were taken, the latter two systolic and diastolic measurements were averaged and used in analyses. If two measurements were taken, the second systolic and diastolic measurement were used in analyses. We then calculated the change in systolic and diastolic BP, considered independently, from enrolment to each postpartum visit.

### Covariates

Data on maternal ethnicity (categorical variable) and age (continuous variable in completed years) were collected through questionnaires during the GRAPHS enrolment visit. The household asset index, a continuous variable to compare the relative wealth of one study household compared to another, was generated from the count enumeration of household assets at GRAPHS enrolment as previously described(32). The child’s biological sex was determined at birth (categorical variable, male/female). Maternal height (in centimeters) and weight (in kilograms) was measured once at enrolment and body mass index (BMI) was calculated. At the four year postpartum visit, and prior to resting BP measurement, we measured HAP exposure via 24-hour personal PM_2.5_ measurement in micrograms/m^3^ using MicroPEM, (RTI International) as previously described (27).

### Statistical Analyses

#### Intention-to-treat Analyses

We examined differences in post-intervention PM_2.5_ exposure by study arm amongst all women with valid four-year postpartum PM_2.5_ exposure measurements using a clustered Wilcoxon rank sum test. Leveraging the longitudinal, repeated BP measurements, we estimated unadjusted and adjusted intention-to-treat linear mixed effects models to examine whether GRAPHS intervention arm (LPG or improved biomass as compared to control) was associated with change in postpartum maternal BP from enrolment through 8 years postpartum. Multivariable regressions adjusted for maternal age at GRAPHS study enrolment and household asset index. To examine the effect over time, we truncated the BP data to use the same model to examine the effect on change in BP at 5 (including 2, 4, 5 year data); 6 (including 2, 4, 5, 6); 7 (including 2, 4, 5, 6, 7) and 8 (including 2, 4, 5, 6, 7, 8 year data) years postpartum. We then explored whether the effect of intervention on maternal BP differed by fetal sex. To do so, we first introduced a study arm (LPG, improved biomass, control) x sex interaction term in the main regression models and then performed sex-stratified regression models. All models included cluster-robust standard errors to account for the cluster randomized design of GRAPHS.

#### Exposure-Response Analyses

To examine the effect of HAP exposure during the GRAPHS intervention period on postpartum BP, we estimated bivariate and multivariable linear mixed effects models to examine associations between average prenatal and postnatal (year 1 postpartum) CO exposure and change in maternal BP from enrolment through 8 years of follow up. Multivariable models adjusted for BP on enrolment, age, ethnicity and body mass index (BMI) on enrolment, and household asset index. A sensitivity model additionally adjusted for PM_2.5_ personal exposure measurement at 4 years postpartum and included propensity score weights to account for missing exposure data. We then explored whether the effect of prenatal and first year postnatal CO exposure different by fetal sex by first introducing an interaction term into the model and second by performed fetal sex stratified analyses.

## Results

Baseline characteristics are shown in table 1. Study participants (n=690) were a median 28 (interquartile range, IQR, 23, 34) years of age on study enrolment and 301, 127 and 262 were assigned to the control, improved biomass and LPG GRAPHS study arms, respectively. Body mass index on enrolment was similar across study arms. Median systolic BP on enrolment was 104mmHg (IQR 99, 110), 104mmHg (IQR 98, 110) and 106mmHg (IQR 100, 115) for control, improved biomass and LPG arms, respectively. Median diastolic BP on enrolment was 62mmHg (IQR 58, 67), 62mmHg (IQR 57, 67) and 63mmHg (IQR 60, 69) for control, improved biomass and LPG arms, respectively. At four years postpartum, amongst all participants who had valid four year postpartum PM_2.5_ measurements, there was no difference in exposure by study arm [24-hour media (IQR): control 64µg/m^3^ (41, 110), improved biomass 61µg/m^3^ (43, 94), LPG 55µg/m^3^ (30, 93), clustered Wilcoxon rank sum p=0.54]. Cohort characteristics of GRAPHS participants included in the present analyses and those not included are presented in **Supplemental Table 1**. Given our sampling framework to overrecruit participants from the LPG arm as opposed to the improved biomass arm, which did not demonstrate exposure reductions(23), there are expected differences in study arm proportions and lower CO exposures in those included versus not included in longitudinal follow up.

**Table 1:**
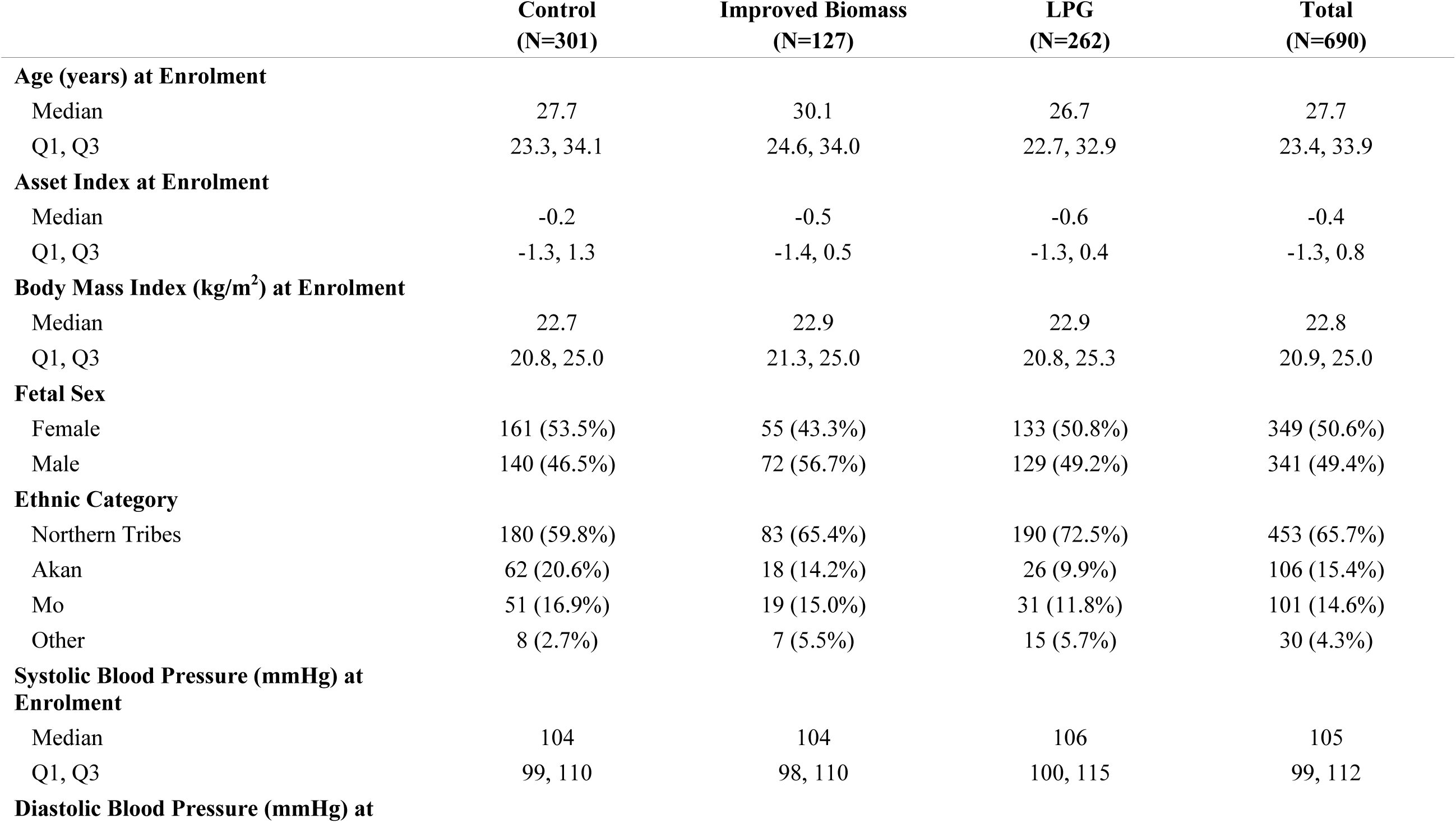
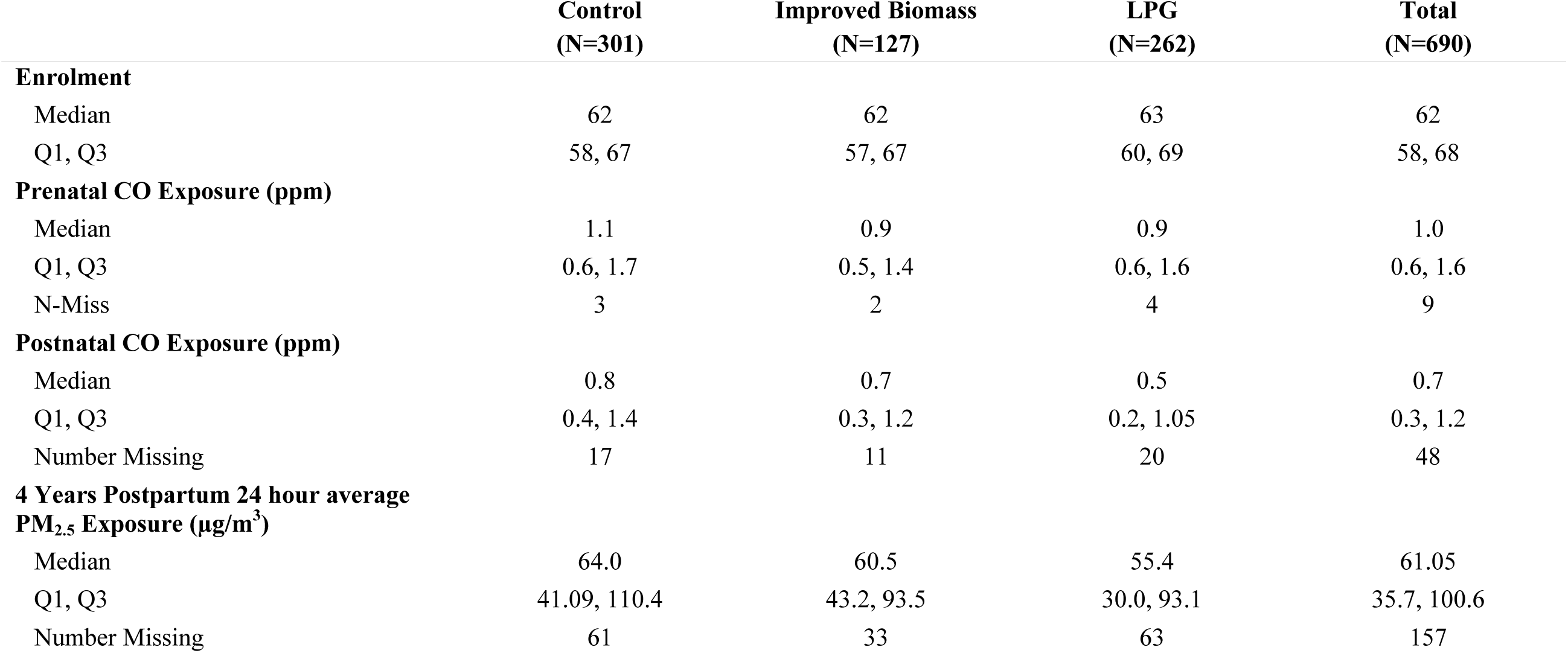
Characteristics of individuals in analysis by study arm.

### Associations between GRAPHS intervention arm and BP change from enrollment

Intention-to-treat models find that on average persons in the LPG arm had 3.54mmHg (95% CI-5.55,-1.53) lower change in systolic BP from enrolment through 8 years postpartum and 2.27mmHg (95% CI-3.61,-0.93) lower change in diastolic BP from enrolment through 8 years postpartum, as compared to persons in the control arm. Adjusting for participant age and household asset index on study enrolment did not substantively change these findings (**Figure 1, Supplemental Table 2**). We did not observe differences in the association by fetal sex (Supplemental Table 2, Supplemental Figure 1).

**Figure 1.**
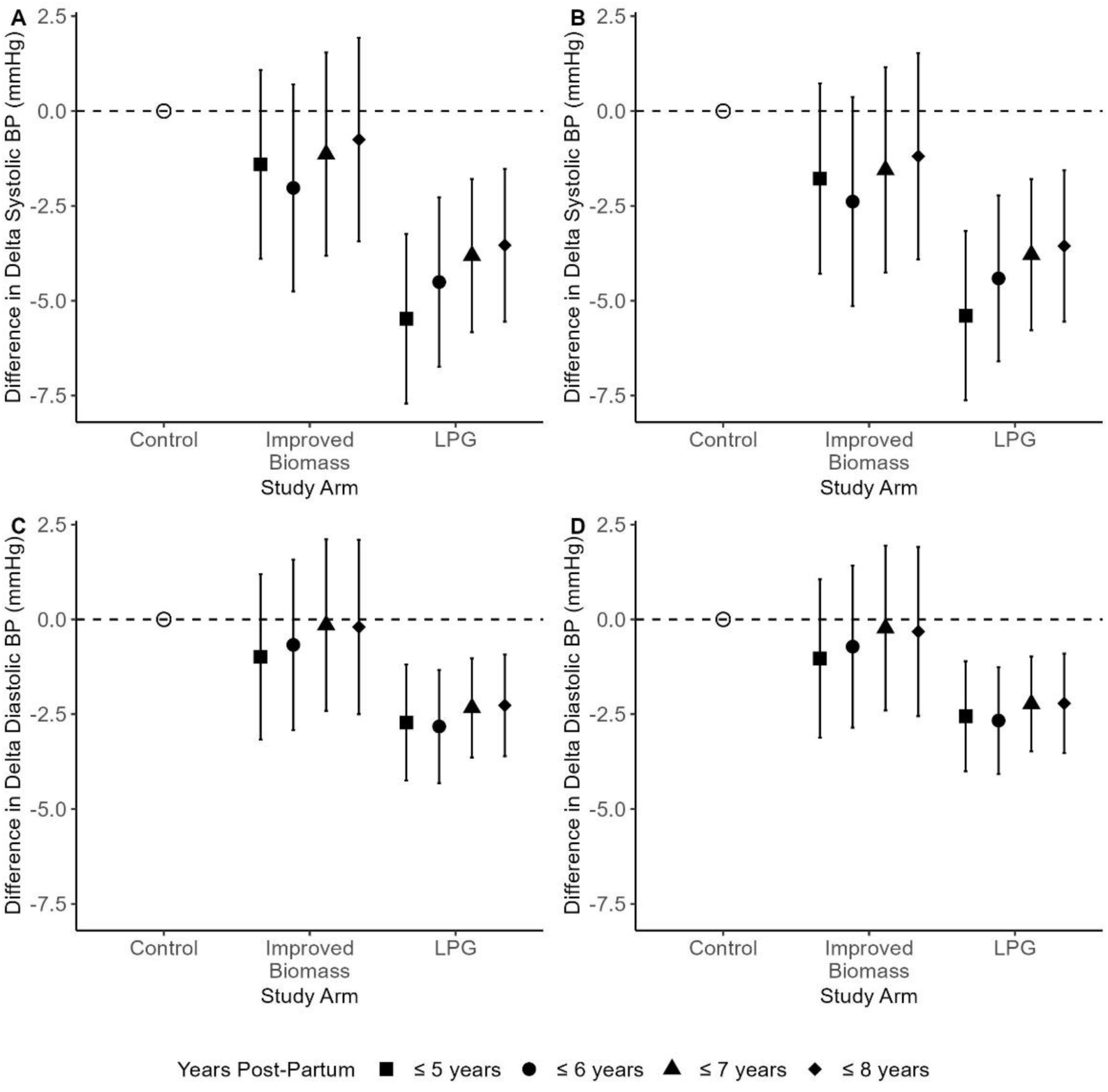
Intention to Treat. Difference compared to the control group in delta blood pressure (BP) from enrollment, through 8 years postpartum. Adjusted models include age and asset index on enrollment. A: Unadjusted delta systolic BP. B: Adjusted delta systolic BP. C: Unadjusted delta diastolic BP. D: Adjusted delta diastolic BP.

The association between study arm and change in systolic BP from enrolment appeared to wane over time. For example, by 5 years postpartum the change in systolic BP from enrolment was 5.48mmHg (95% CI-7.71,-3.24) lower in the LPG arm as compared to control while by 7 years the change in systolic BP from enrolment was 3.81mmHg (-5.83,-1.79) lower in the LPG arm as compared to control (**Figure 1, Supplemental Table 3**). Conversely, the associations between study arm and change in diastolic BP from enrolment appear relatively stable over time. For example, by 5 years postpartum the change in diastolic BP from enrolment was 2.72mmHg (95% CI-4.25,-1.19) lower in the LPG arm as compared to control and at 7 years the change in diastolic BP from enrolment was 2.33mmHg (95% CI-3.64,- 1.03) lower in the LPG arm as compared to control (**Figure 1, Supplemental Table 3**). As with the overall intention-to-treat association, these associations did not appear to differ by fetal sex (**Supplemental Table 3, Supplemental Figure 1**).

### Associations between average prenatal and first year postpartum HAP exposure and BP change from enrolment

Average prenatal and first year postpartum CO exposures were weakly correlated (Spearman correlation rho = 0.10, p-value = 0.01) and n=637 participants had both prenatal and first year postpartum CO measures. Multivariable models adjusting for participant age, BP, and BMI on enrolment, and ethnicity and household asset index did not observe associations between prenatal or postnatal CO exposure and systolic or diastolic BP, considered separately (**Table 2**). Models stratified by fetal sex, identified that amongst pregnant persons carrying a male fetus, prenatal average CO exposure was positively associated with change in systolic BP from enrolment (β=0.71mmHg, 95% CI 0.08, 1.30, per doubling of CO) and a trend was observed with change in diastolic BP from enrolment (β=0.40mmHg, 95% CI-0.04, 0.84, per doubling of CO).

**Table 2.**

| Delta BP Measurement | N | # Obs | Multivariable Model<sup>e</sup> |  |  |  | N | # Obs | Multivariable Model (Sensitivity)<sup>c, d</sup> |  |  |  |
| --- | --- | --- | --- | --- | --- | --- | --- | --- | --- | --- | --- | --- |
|  |  |  | Prenatal |  | Postnatal |  |  |  | Prenatal |  | Postnatal |  |
| | | | $\beta$ (95% CI<sup>f</sup>) | p | $\beta$ (95% CI<sup>f</sup>) | p | | | $\beta$ (95% CI<sup>f</sup>) | p | $\beta$ (95% CI<sup>f</sup>) | p |
| All pregnancies |  |  |  |  |  |  |  |  |  |  |  |  |
| SBP | 637 | 2954 | 0.25 (-0.20, 0.70) | 0.29 | 0.14 (-0.17, 0.43) | 0.36 | 493 | 2308 | 0.46 (-0.12, 1.04) | 0.093 | 0.16 (-0.21, 0.48) | 0.35 |
| DBP | 637 | 2954 | 0.13 (-0.19, 0.45) | 0.46 | 0.09 (-0.13, 0.30) | 0.40 | 493 | 2308 | 0.18 (-0.23, 0.61) | 0.36 | 0.11 (-0.14, 0.35) | 0.35 |
| Male pregnancy<sup>g, h</sup> |  |  |  |  |  |  |  |  |  |  |  |  |
| SBP | 317 | 1464 | **0.71 (0.08, 1.30)** | **0.02** | 0.11 (-0.25, 0.50) | 0.57 | 242 | 1127 | **0.93 (0.21, 1.62)** | **< 0.01** | 0.27 (-0.19, 0.81) | 0.22 |
| DBP | 317 | 1464 | 0.40 (-0.04, 0.84) | 0.07 | 0.11 (-0.15, 0.39) | 0.45 | 242 | 1127 | **0.48 (-0.05, 1.00)** | **0.053** | 0.15 (-0.18, 0.55) | 0.34 |
| Female pregnancy<sup>g, h</sup> |  |  |  |  |  |  |  |  |  |  |  |  |
| SBP | 320 | 1490 | -0.33 (-1.03, 0.42) | 0.36 | 0.16 (-0.28, 0.55) | 0.48 | 251 | 1181 | -0.19 (-1.17, 0.75) | 0.67 | 0.06 (-0.50, 0.60) | 0.81 |
| DBP | 320 | 1490 | -0.22 (-0.73, 0.32) | 0.40 | 0.06 (-0.26, 0.36) | 0.72 | 251 | 1181 | -0.24 (-0.96, 0.43) | 0.46 | 0.07 (-0.33, 0.47) | 0.71 |

<sup>a</sup> CO measured by repeated personal exposure measurements and time-weighted average over pregnancy and the first year postpartum. Log base 2 transformed, so resulting interpretation is change in delta BP per doubling of CO.
<sup>b</sup> Mixed effects models examining associations between household air pollution exposure over pregnancy and postnatal period and change in postnatal blood pressure from enrolment trajectories through eight years postpartum.
<sup>c</sup> Sensitivity analysis model has the same covariates as the multivariable model<sup>e</sup>, with an additional adjustment for 24-hour average PM2.5 exposure measured four years post-partum (log base 2 transformed).
<sup>d</sup> Propensity score of having year 4 PM2.5 pollution exposure measurement calculated using logistic regression with the following covariates at enrolment: blood pressure (systolic or diastolic, depending on model outcome), age, asset index, body mass index, and ethnicity.
<sup>e</sup> Multivariable models adjust for blood pressure (systolic or diastolic, depending on model outcome) on enrolment, age at enrolment, asset index, body mass index on enrolment and ethnicity.
<sup>f</sup> Bootstrap percentile confidence intervals.
<sup>g</sup> For the delta SBP outcome multivariable model, the p-values of the interaction terms between fetal sex and prenatal exposure and fetal sex and postnatal exposure are 0.079 and 0.85 respectively. For the delta DBP outcome multivariable model, the p-values of the interaction terms between fetal sex and prenatal exposure and fetal sex and postnatal exposure are 0.18 and 0.87 respectively.
<sup>h</sup> For the delta SBP outcome multivariable sensitivity model, the p-values of the interaction terms between fetal sex and prenatal exposure and fetal sex and postnatal exposure are 0.13 and 0.56 respectively. For the delta DBP outcome multivariable sensitivity model, the p-values of the interaction terms between fetal sex and prenatal exposure and fetal sex and postnatal exposure are 0.20 and 0.74 respectively.

Sensitivity models additionally adjusting for PM2.5 exposure at 4 years postpartum, representing post-intervention exposure, identified a trend in effect of prenatal CO on change in systolic BP from enrolment in the overall group (β=0.46mmHg, 95% CI-0.12, 1.04 per doubling of CO) and associations between prenatal CO and change in systolic BP (β=0.93mmHg, 95% CI 0.21, 1.62 per doubling of CO) and diastolic BP (β=0.48, 95% CI-0.05, 1.00) in pregnant persons carrying a male fetus (**Table 2**).

## Discussion

Pregnancy is a critical window where future CVD risk may be programmed. Despite the billions of people worldwide that are exposed daily to HAP, including many pregnant persons, very little is known about associations between HAP exposure over pregnancy and the immediate postpartum period and longer-term postpartum CVD risk. The data presented herein from the GRAPHS cohort with 8 years of longitudinal postpartum BP follow up find that a cleaner fuel stove intervention, initiated during pregnancy and previously associated with a 47% reduction in CO exposure, is associated with better systolic and diastolic BP trajectories through 8 years postpartum, as compared to control. These data suggest a sustained CVD risk benefit. Given the importance of BP in determining future CVD risk, this finding has broad public health significance.

Exposure-response analyses find that higher prenatal HAP exposure especially amongst persons pregnant with a male fetus is associated with higher BP trajectories. Taken together, these findings highlight the importance of air pollution on CVD risk and suggest that the prenatal period may be critical windows of susceptibility for programming subsequent CVD risk. Further, these findings suggest that a prenatally delivered stove intervention to reduce air pollution exposures among biomass fuel users may be an effective public health intervention to reduces postpartum BP trajectories, with implications for life course CVD risk.

Prior air pollution epidemiological studies find that higher ambient air pollution exposures are associated with higher risk of adverse pregnancy outcomes (33–35), including adverse pregnancy outcomes linked to subsequent CVD risk such as small-for-gestational age (SGA) or preterm birth (PTB)(36–38). Indeed, prior studies have also identified associations between gestational air pollution exposure and hypertensive disorders of pregnancy (17–19). Fewer studies have examined HAP health sensitivities, however some, including GRAPHS, similarly find that higher HAP exposure over pregnancy is associated with higher risk for SGA babies or PTB(28, 33, 39, 40). Few studies have extended these findings beyond the immediate postpartum period to assess the impact of environmental exposures over pregnancy on longitudinal CVD risk. We present evidence in the context of a cluster randomized controlled trial with demonstrated HAP exposure reductions over pregnancy and postpartum periods that find that HAP exposure over pregnancy and the first year postpartum is important in shaping postpartum BP trajectories, a central component of cardiovascular health. Exposure-response analyses point to the importance of the prenatal period.

Previous studies on the effects of HAP exposure on maternal cardiovascular health have focused on gestational blood pressure, and have shown that higher exposures to HAP constituents such as PM2.5 and CO are associated with a higher risk of developing HDP like gestational hypertension and pre-eclampsia/eclampsia(20, 41). Studies examining the long-term maternal cardiovascular health outcomes post-pregnancy have done so mainly in relation to previous history of HDP. The effects of HAP exposure in the prenatal and or early postnatal periods on maternal post-pregnancy BP trajectories are unknown.

Profound hemodynamic changes in pregnancy increase the risk of HDP and postpartum CV risk (42, 43) thus making pregnancy a sensitive period for CVD development. Additional insults from environmental exposures may exacerbate these risks. Exposure to air pollution may promote CVD progression via multifactorial mechanisms(44, 45), and we posit that these mechanisms may be amplified during pregnancy. We hypothesize that the air pollution exposure response over pregnancy, through well-described pathways of inflammation and oxidative stress(46–48) – mechanisms also central to CVD development – are pathways that are already altered over the course of pregnancy. We thus hypothesize that the converging pathways results in a sensitive window of exposure for the pregnant person. Inhalation of HAP constituents causes lung and systemic inflammation and oxidative damage which may induce endothelial dysfunction and vascular inflammation(49).

PM_2.5_ triggers a T-reg and Th17 imbalance(50), which animal models suggest has a central role in air pollution-induced preeclampsia(51).

Well-established literature links Hypertensive Disorders of Pregnancy (HDP) and, specifically, Preeclampsia (PE) to long-term CVD risks, including BP (52–57). At the core of HDP/Preeclampsia pathology is the placental dysfunction resulting from placental vascular abnormalities, which can, in turn, induce changes to the maternal vascular system, with long-lasting CV consequences(58, 59). Pathways and sources of susceptibility for placental dysfunction in HDP are still unclear. Previous studies have linked air pollution, including HAP to increased inflammation, oxidative stress, and endothelial dysfunction(60–63), which limits effective placental circulation and oxygenation, and may influence the development of placental dysfunction and HDP. Further, maternal HAP exposure has been associated with thrombotic placental lesions(64) and abnormal vascularization(65) that impact placental function. It is possible that HDP and maternal CVD later in life represent the manifestations of these multifaceted pathophysiological mechanisms. Further, evidence is emerging of a significant incidence of postpartum hypertension (PPHTN) following normotensive pregnancy suggested that the complete manifestation of HDP may not be necessary. A recent retrospective study at a safety-net hospital in Boston, Massachusetts estimated a 12.1% incidence of hypertension at one year postpartum among a cohort of 2465 women without a history of chronic hypertension or HDP(66). In a sub-analysis, the authors also found approximately 33% risk of de novo PPHTN in ≥ 35-year-olds with caesarean delivery and any tobacco smoking exposure (current or former).

There is little data on effective environmental interventions to improve BP in sub-Saharan Africa, despite the high prevalence of HTN(67). With the LPG stove intervention, we observe that, on average, women in the LPG arm had 3.5mmHg lower change in systolic BP and a 2.3mmHg lower change in diastolic BP than women in the Control arm over 8 years of postpartum follow up. The effect size for systolic BP in this study is similar in magnitude to that found in a systematic review and meta-analysis of interventions targeting dietary salt intake and lifestyle modifications in Sub-Saharan Africa (68). For diastolic BP, these effect sizes are similar in magnitude to those identified in a systematic review and meta-analysis of dietary interventions in adults(69). On a population level, the Framingham Heart Study and the National Health and Nutrition Examination Survey II identified that a 2mmHg reduction in diastolic BP would translate to 15% lower risk of stroke and transient ischemic attacks, 6% lower risk of coronary heart disease, and a 17% reduction in the prevalence of hypertension(70). Similarly, a 5mmHg reduction in systolic BP would translate to a 10% risk reduction of major cardiovascular events(71). We hypothesize that these population-level effects may be even more pronounced in areas where there is limited capacity for diagnostic and preventative care. Thus, public policy promoting clean energy transitions may deliver significant CVD benefits.

Our study adds to the limited body of evidence examining associations between HAP exposure over pregnancy and postpartum BP trajectories. Strengths of this study include a cluster RCT design with a randomized air pollution differential over pregnancy – a hypothesized sensitive window of exposure -- and longitudinal BP follow up through 8 years postpartum. Repeated personal exposure assessments over pregnancy and postpartum allow for exposure-response analyses, in addition to intention-to-treat findings. We characterize BP trajectories with up to 6 annual postpartum measurements, each measurement performed in duplicate or triplicate, per participant. The well-characterized GRAPHS cohort allows for adjustment for individual-and household-level covariates. We also note limitations. BP measurements occurred primarily in the morning although we did not record the exact time of BP measurement, nor activity or diet immediately prior to measurement. We did not record BP on successive days. We hypothesize that any bias introduced by these factors would be non-differential and bias towards the null.

## Conclusion

In summary, we find that pregnant study participants randomized to an LPG stove intervention over pregnancy and the first year postpartum had lower systolic and diastolic BP trajectories over 8 years postpartum. Exposure response findings support the importance of HAP exposures during the prenatal period. These findings support the need for improved air quality, especially over pregnancy.

### Perspectives

Pregnancy is a sensitive window during which lifetime CVD risk may become established. As the primary cook in most LMIC settings, women are highly exposed to HAP including during pregnancy. Evidence linking HAP exposure in pregnancy to BP is scarce and limited to BP measurements taken during pregnancy. Leveraging longitudinal data collected up to 8 years postpartum in the context of a randomized controlled cookstove intervention trial in Ghana, we provide new evidence on the protective effect of a clean-burning LPG cookstove intervention to reduce HAP on BP. LPG intervention over pregnancy to the first year postpartum was associated with lower systolic and diastolic BP up to 8 years postpartum. Exposure-response analysis suggested that higher prenatal HAP exposures, as indexed by CO, was associated with higher systolic and diastolic BP. These findings underscore the role of prenatal HAP exposures on higher postpartum BP and the importance of intervening early in pregnancy to protect women’s cardiovascular health.

### Novelty and Relevance

- This is the first study of its kind to link randomized stove assignment to longitudinal postpartum blood pressure
- LPG cookstove intervention over pregnancy and the first postpartum year reduced systolic and diastolic BP trajectories up to 8 years postpartum
- Clean cooking solutions to reduce HAP exposure could be a relevant public health intervention to reduce BP at the population level and contribute to the primary prevention of hypertension in women.

## Non-standard Abbreviations and Acronyms

CO: Carbon Monoxide
DALY: Disability Adjusted Life Years
GRAPHS: Ghana Randomized Air Pollution and Health Study
HAP: Household Air Pollution
HAPIN: Household Air Pollution Intervention Network
HDP: Hypertensive Disorders of Pregnancy
KHRC: Kintampo Health Research Centre
LPG: Liquefied Petroleum Gas
PM2.5: Particulate Matter with aerodynamic diameter <2.5 microns
PE: Preeclampsia
PTB: Preterm birth
PPHTN: Postpartum hypertension
SGA: Small-for-gestational age

## Data Availability

Anonymized data that underlie the results reported herein are available upon request. Proposals should be directed to kwakupoku. and to; to gain access, data requestors will need to sign a data access agreement

## Acknowledgements

The authors acknowledge the community leaders in the study area, study participants and study staff, and the Ghana Health Service facilities in the Kintampo North Municipality and Kintampo South District.

## Disclosures

All authors declare that they have no known competing financial interests or personal relationships that could have appeared to influence the work reported in this paper.

The results, conclusions and recommendations in this paper are those of the authors and do not represent the official position US National Institutes of Health.

Change in Maternal Blood Pressure from Enrolment - Intention to Treat and Exposure-Response

